# Awareness of cervical cancer and screening in Benin and Cameroon: analysis of the Demographic and Health Survey (DHS)

**DOI:** 10.1101/2023.01.03.23284133

**Authors:** Nike Olajide, Bhautesh Dinesh Jani, Claire L Niedzwiedz, Cathy Johnman, Kathryn A. Robb

## Abstract

Cervical cancer incidence and mortality are high in Africa. We assessed cervical cancer and screening awareness in two West African countries.

We used data from the Demographic and Health Survey (DHS) in Benin (DHS 2017-2018) and Cameroon (DHS 2018). Women (n=21322) aged 15-49 were interviewed on awareness of cervical cancer and cervical cancer screening. Descriptive statistics and logistic regression analysis were used.

Awareness of cervical cancer was low among women in Cameroon (46.1%) and very low in Benin (9.5%). Among those aware of cervical cancer, 51.4% in Benin and 59.7% in Cameroon were also aware of cervical screening. In the adjusted analysis, women in Cameroon aged 45+ had the highest awareness odds of both cervical cancer (adjusted odds ratio-aOR 2.91, [2.36-3.60]) and screening (aOR 1.75, [1.33-2.29]). In Benin, women aged 45+ had the highest cervical cancer awareness (aOR 1.89 [1.23-2.91]) while screening awareness was highest in women aged 25-34 years (aOR 1.98, [1.20-3.27]). Women with higher education were six to nine times more aware of cervical cancer and three to four times more aware of cervical screening than women with no education in Benin and Cameroon respectively. Additionally, cervical cancer awareness was approximately four times higher in the richest wealth quintile in Cameroon. In Benin, the odds of awareness of cervical cancer were increased with daily internet use (aOR 3.61, [2.45-5.32]) and radio listening once a week (aOR 1.73 [1.04-2.88]) compared to no internet and no radio listening respectively. In Cameroon, both awareness of cervical cancer and screening increased with daily internet use (aOR 1.95 [1.61-2.35]) and (aOR 1.35 [1.10-1.67]) respectively.

There is a need to increase awareness of cervical cancer and screening in Benin and Cameroon and likely among other West African countries. The internet and radio appear to be important potentially effective means for raising awareness.

## Introduction

The global incidence rate of cancer has risen from 12.7 million in 2008 to 19.3 million in 2020 (1, 2). Cancer causes significant morbidity and mortality worldwide, regardless of socioeconomic development (2). The majority of the deaths from cervical cancer occur in Sub-Saharan Africa, Melanesia, South America, and South-Eastern Asia (2-5). The age-standardised incidence rates ASR (25.6 per 100,000 population) and the cumulative risk (3.86%) of cervical cancer is highest in Africa compared to other continents (6). In 2020, the ASR per 100 000 women in Benin and Cameroon were 15.1 and 33.7 per 100 000 women respectively (7, 8), and the cumulative risk of cervical cancer in ages 0-74 in both countries was 1.8% and 3.6% respectively (7, 8). In 2018, cervical cancers attributable to Human Papilloma Virus (HPV) 16 and 18 accounted for an estimated 72% of cases globally (9) (10).

Cervical cancer can be prevented or detected early using the HPV vaccine and screening strategies. Indeed, the World Health Organisation (WHO) has a global strategy to eliminate cervical cancer (11). WHO’s target age for vaccination in girls is between the ages of 9-14 years, before the onset of sexual intercourse (12). A systematic review revealed that between 2006 and 2014, only 1% of Low Middle-Income Countries (LMICs) had HPV vaccination programmes (13). Screening is recommended using cervical cytology alone every three years for women aged 21 to 29 years (14). For women aged 30 to 65 years, three-yearly cytology, five-yearly with high-risk human papillomavirus (hrHPV) testing, or both (co-testing) five yearly is recommended (14). Due to the difficulty in implementing screening programmes because of poor infrastructures and funding in low-resource settings like Africa, the WHO recommends a screen-and-treat approach to allow for treatment based on the result of the screening test (15). Most African countries use visual inspection with acetic acid (VIA) and immediate treatment with cryotherapy, while some continue with the use of cytology-based screening or are examining HPV testing (15, 16).

Lifetime uptake data of cervical cancer screening in 55 LMICs countries (representing 72% of the world population), reveals that only 44% of women in the target age group have ever had the opportunity to get a cervical screening test, with values even lower in sub-Saharan Africa (country level median 16.9%; range 0.9% - 50.8%) in contrast to more than 60% in High-Income Countries (17, 18). WHO enjoins introducing HPV vaccination and expanding screening and treatment for effective cervical cancer prevention (19, 20). Although cervical cancer incidence and mortality are high in low-resource settings including Africa, several potentially modifiable factors are implicated including a lack of awareness of the symptoms, and screening (21), poor communication, and inadequate media education (22). The present study explored levels of awareness of cervical cancer and cervical screening in two West African countries, Benin, and Cameroon. Awareness is an important first step in increasing engagement in screening and identifying characteristics of those with lower levels of awareness can assist in the development of targeted interventions. This study addressed the following research questions:

1. What is the level of awareness of cervical cancer in Benin and Cameroon?
2. What is the level of awareness of cervical screening in Benin and Cameroon?
3. Does the level of awareness of cervical cancer and cervical screening differ by demographic characteristics and engagement with media?

## Materials and Methods

### Study design and setting

This study uses the Demographic and Health Survey (DHS) of two West African countries (Benin and Cameroon) conducted between 2017-2018 (Fig 1). Eighteen West African countries have data in the DHS but only two collected data on cervical cancer and cervical screening between 2017-2020 (http://dhsprogram.com/).

**Fig 1.**
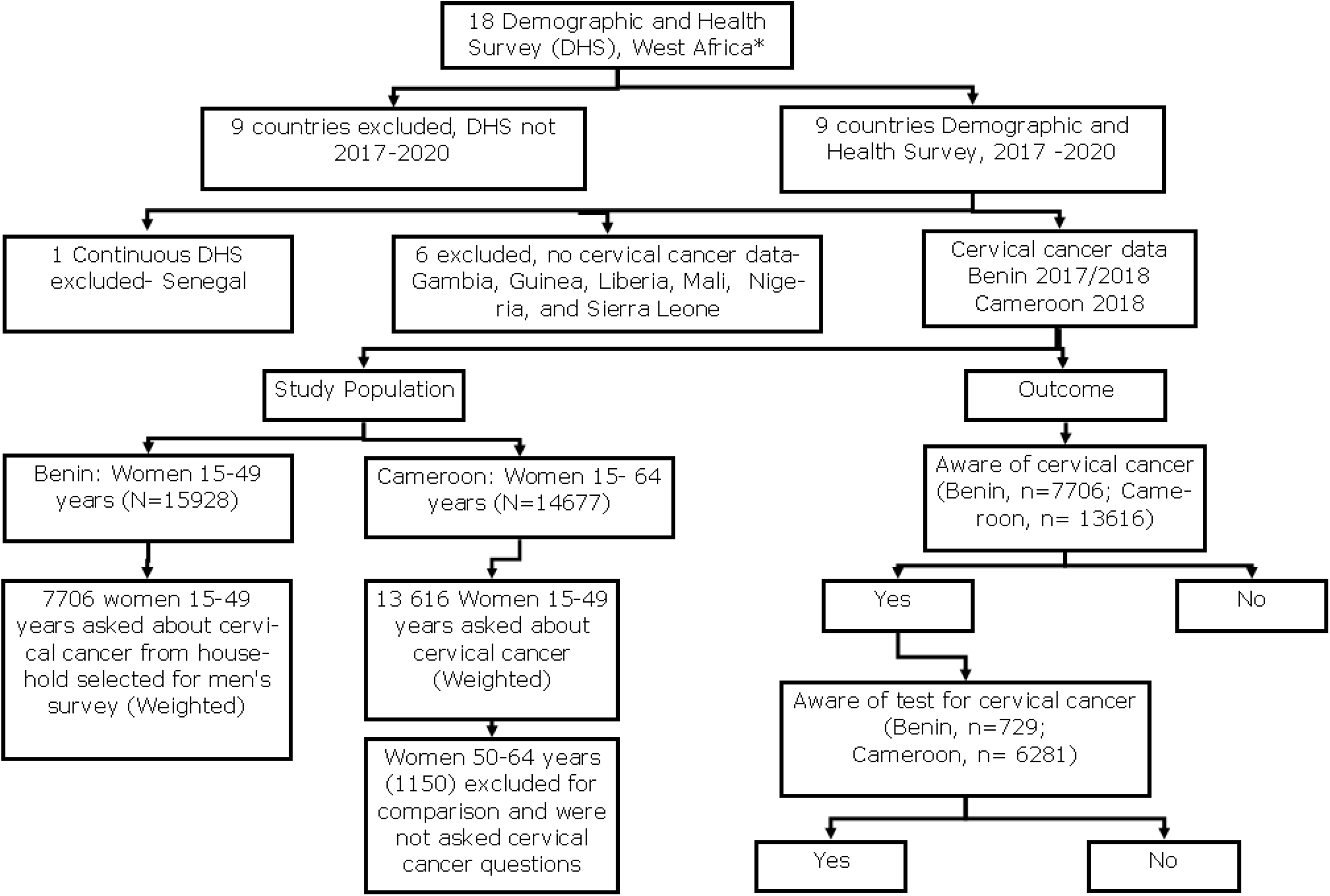
Flow chart of sample selection. *https://www.britannica.com/place/western-Africa

### Study population

Fig 1 shows a flow chart of the sample selection process. Cameroonian women aged 50-64 years were excluded because they were not interviewed about cervical cancer.

### Measures

#### Awareness of cervical cancer and screening

The outcomes of interest were whether women were aware of cervical cancer and cervical screening. The questionnaire was written in French. In both countries, the women were asked “Have you heard about cervical cancer?” and if ‘Yes’, they were then asked, “Have you heard of testing for cervical cancer of the uterus?” (https://translate.google.co.uk/). The responses were either ‘Yes’ or ‘No’ in both outcomes (Fig 1).

#### Demographic characteristics

Age was measured in 10-year age group intervals from 15 – 49 years. Residential status was assessed as ‘urban’ or ‘rural’. For marital status, ‘never married’ were women who had never lived with a partner, the ‘currently married’ had married or were not formally married but were living with a partner, and the ‘formerly married’ women were widowed, divorced, or separated, or who had formerly lived with a partner. We grouped marital status as ‘never in a union’ and ‘currently/ formerly in a union’. Education assessed the highest achieved level and was grouped into four categories (no education, primary, secondary, higher) in line with the standard DHS report. The wealth quintile was measured as the relative wealth of the household based on ownership of assets such as radio, television, car, motorcycle, and materials for housing construction (23), and was divided into quintiles of poorest, poorer, middle, higher, and highest. The categories of religion were country-specific, but were broadly categorized as ‘Christians’, ‘Muslims’, ‘none’, and ‘others’. Occupation of the women were reported based on the standardized DHS format but were recategorized as ‘not working’, ‘non-professionals’, and ‘professionals’.

#### Media use

The frequency of reading the newspaper, watching television, listening to the radio, and using the internet in the last month was reported as, ‘not at all’, ‘less than once a week’, and ‘at least once a week’. For frequency of using the internet in the last month, women who use it almost every day were also included.

### Ethics Considerations

We obtained approval to use the data from the DHS program. The DHS program has ethical documentation on the privacy and confidentiality of all the respondents (http://dhsprogram.com/).

### Statistical Analysis

Univariate descriptive statistics were used to describe the demographic characteristics and media use. We summarised the categorical variables as weighted frequencies and percentages while the numerical variables were summarised as weighted means with a 95% Confidence Interval (CI). A Chi-square test was used to determine the association between demographic characteristics and the awareness of cervical cancer/screening. All variables were included in the multivariate analysis. Multivariate logistic regression was then used to determine the association between demographic characteristics and media use and awareness of cervical cancer and screening. The association was reported as odds ratio (OR), P values, and 95% CI.

We used the survey package in R version 4.1.1 (2021-08-10), to handle the survey design (24). Each country was analysed separately, and we applied sample weights for all the descriptive statistical analyses. The regression analysis considered the survey design by using the primary sampling unit (PSU), secondary sampling unit (SSU), sample strata, and weight.

## Results

### Demographic characteristics

A total of 21322 women of reproductive age were surveyed in Benin (n=7706) and Cameroon (n=13616) on awareness of cervical cancer and 7010 of these women were additionally asked about their awareness of cervical cancer screening. Table 1 shows the demographic characteristics of the women included in this study. The mean age was slightly lower in Cameroon (27.85 [95% CI 27.66-28.04]) than in Benin (28.39 [95% CI 28.15-28.62]). The majority of the women were within the age group 15 – 24 years in both countries, as high as 42.1% in Cameroon. Most women resided in a rural area, Benin (51.7%) and Cameroon (68.5%). More women in Benin (75.3%) than in Cameroon (54.8%) had either been married or were formerly married. More than half of the women (55.1%) in Benin had no education but 45.2% of women in Cameroon had secondary education. The richest wealth quintile had the highest proportion in Benin (22.5%) and Cameroon (23.8%). Most of the women were Christians, Benin (54.9%) and Cameroon (71.7%). The majority of the women were nonprofessionals, Benin (74.6%) and Cameroon (64.7%).

**Table 1.**
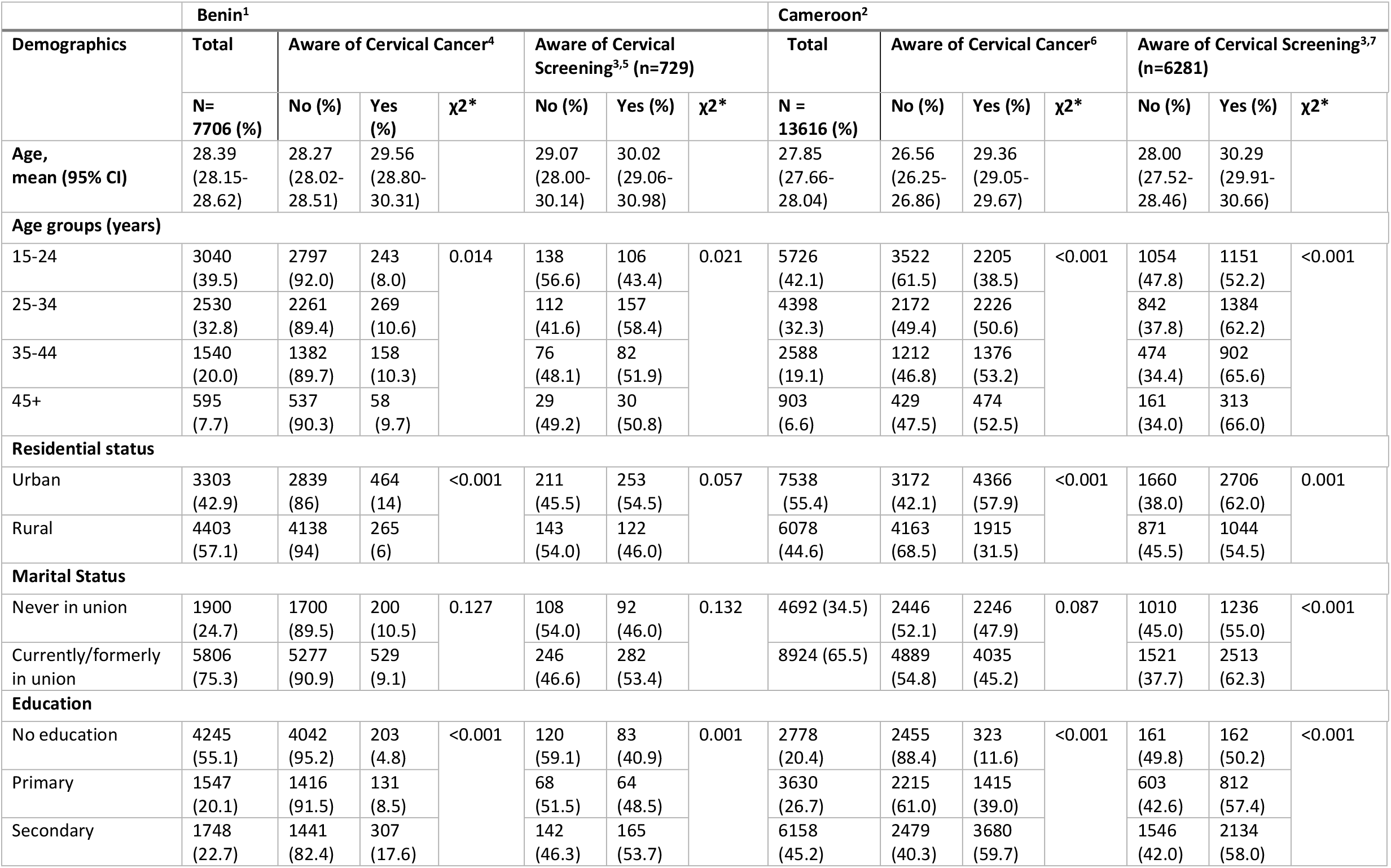
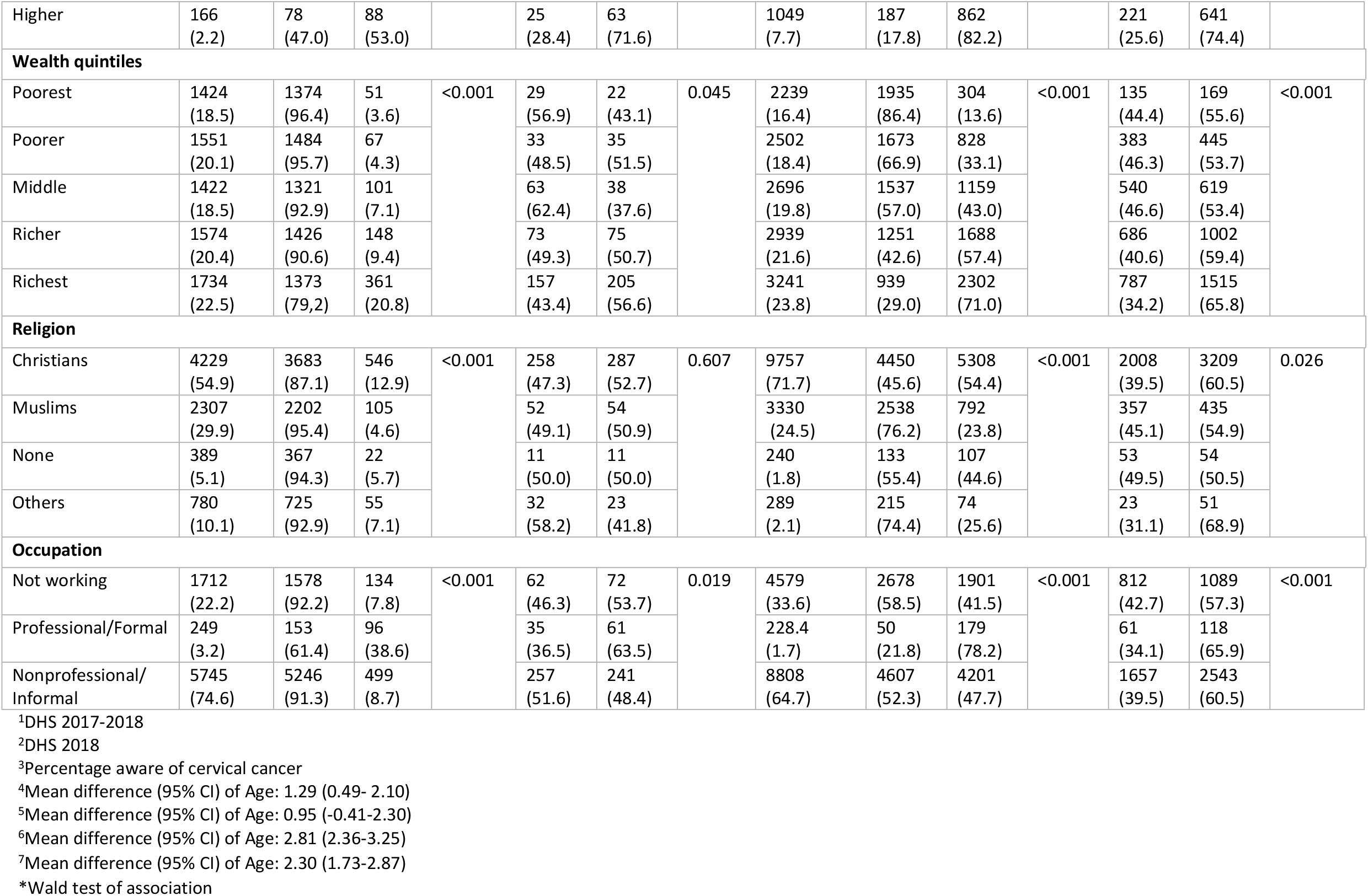
Demographic characteristics and awareness of cervical cancer and screening in women aged 15-49 in Benin and Cameroon (weighted)

### Media use

Most of the women in Benin did not read newspapers/magazines (90.4%; Table 2), watch television (62.5%), listen to the radio (42.8%), or use the internet (93.8%). Similarly, in Cameroon, the majority of the women did not read newspapers/magazines (80.5%), watch television (42.8%), listen to the radio (61.5%), or use the internet (73.5%; Table 2).

**Table 2.**
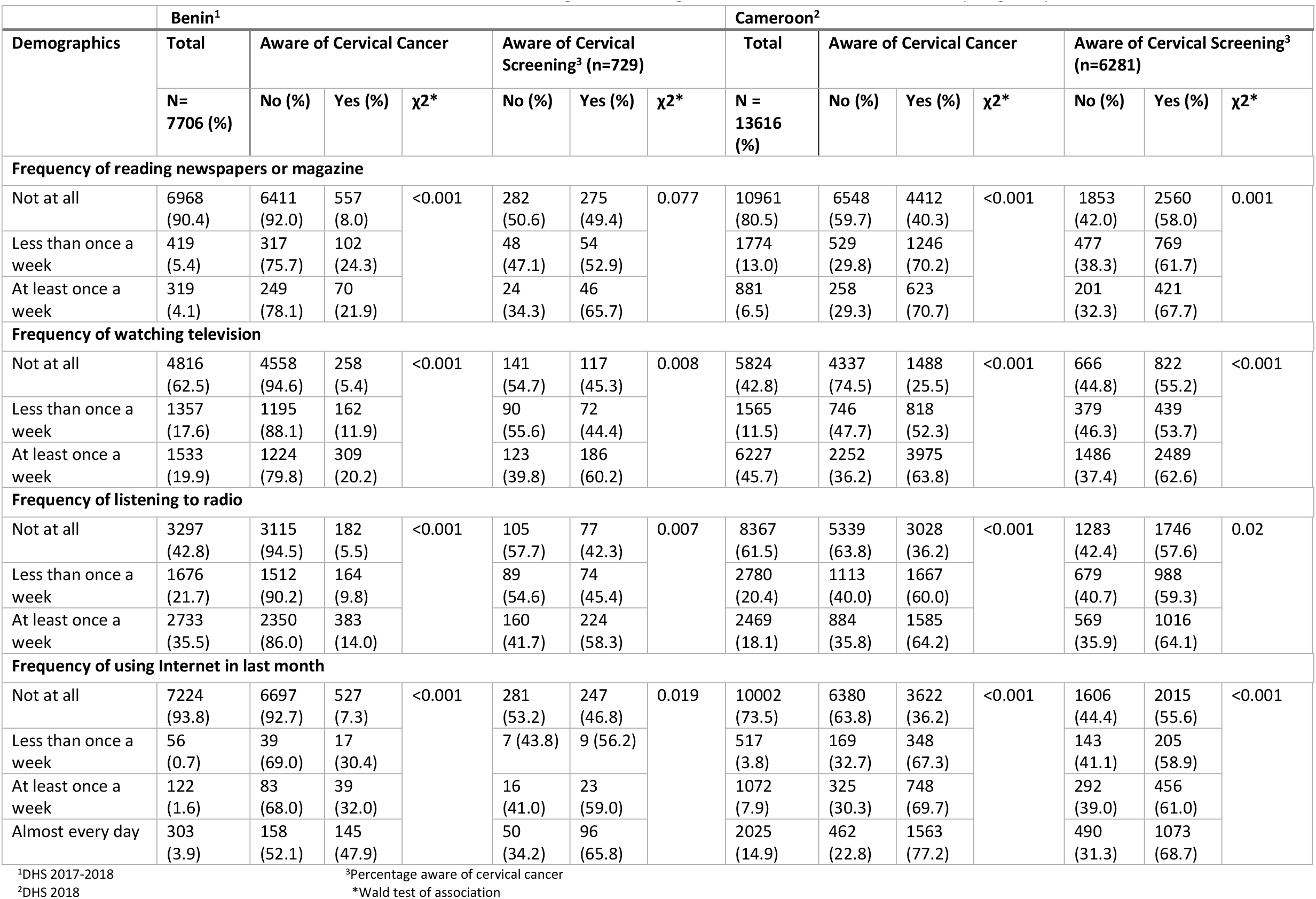
Media use and awareness of cervical cancer and screening in women aged 15-49 in Benin and Cameroon (weighted)

|  | Benin<sup>1</sup> |  |  |  |  |  |  | Cameroon<sup>2</sup> |  |  |  |  |  |  |
| --- | --- | --- | --- | --- | --- | --- | --- | --- | --- | --- | --- | --- | --- | --- |
| Demographics | Total | Aware of Cervical Cancer |  |  | Aware of Cervical Screening<sup>3</sup> (n=729) |  |  | Total | Aware of Cervical Cancer |  |  | Aware of Cervical Screening<sup>3</sup> (n=6281) |  |  |
|  | N= 7706 (%) | No (%) | Yes (%) | χ2* | No (%) | Yes (%) | χ2* | N = 13616 (%) | No (%) | Yes (%) | χ2* | No (%) | Yes (%) | χ2* |
| Frequency of reading newspapers or magazine |  |  |  |  |  |  |  |  |  |  |  |  |  |  |
| Not at all | 6968 (90.4) | 6411 (92.0) | 557 (8.0) | <0.001 | 282 (50.6) | 275 (49.4) | 0.077 | 10961 (80.5) | 6548 (59.7) | 4412 (40.3) | <0.001 | 1853 (42.0) | 2560 (58.0) | 0.001 |
| Less than once a week | 419 (5.4) | 317 (75.7) | 102 (24.3) |  | 48 (47.1) | 54 (52.9) |  | 1774 (13.0) | 529 (29.8) | 1246 (70.2) |  | 477 (38.3) | 769 (61.7) |  |
| At least once a week | 319 (4.1) | 249 (78.1) | 70 (21.9) |  | 24 (34.3) | 46 (65.7) |  | 881 (6.5) | 258 (29.3) | 623 (70.7) |  | 201 (32.3) | 421 (67.7) |  |
| Frequency of watching television |  |  |  |  |  |  |  |  |  |  |  |  |  |  |
| Not at all | 4816 (62.5) | 4558 (94.6) | 258 (5.4) | <0.001 | 141 (54.7) | 117 (45.3) | 0.008 | 5824 (42.8) | 4337 (74.5) | 1488 (25.5) | <0.001 | 666 (44.8) | 822 (55.2) | <0.001 |
| Less than once a week | 1357 (17.6) | 1195 (88.1) | 162 (11.9) |  | 90 (55.6) | 72 (44.4) |  | 1565 (11.5) | 746 (47.7) | 818 (52.3) |  | 379 (46.3) | 439 (53.7) |  |
| At least once a week | 1533 (19.9) | 1224 (79.8) | 309 (20.2) |  | 123 (39.8) | 186 (60.2) |  | 6227 (45.7) | 2252 (36.2) | 3975 (63.8) |  | 1486 (37.4) | 2489 (62.6) |  |
| Frequency of listening to radio |  |  |  |  |  |  |  |  |  |  |  |  |  |  |
| Not at all | 3297 (42.8) | 3115 (94.5) | 182 (5.5) | <0.001 | 105 (57.7) | 77 (42.3) | 0.007 | 8367 (61.5) | 5339 (63.8) | 3028 (36.2) | <0.001 | 1283 (42.4) | 1746 (57.6) | 0.02 |
| Less than once a week | 1676 (21.7) | 1512 (90.2) | 164 (9.8) |  | 89 (54.6) | 74 (45.4) |  | 2780 (20.4) | 1113 (40.0) | 1667 (60.0) |  | 679 (40.7) | 988 (59.3) |  |
| At least once a week | 2733 (35.5) | 2350 (86.0) | 383 (14.0) |  | 160 (41.7) | 224 (58.3) |  | 2469 (18.1) | 884 (35.8) | 1585 (64.2) |  | 569 (35.9) | 1016 (64.1) |  |
| Frequency of using Internet in last month |  |  |  |  |  |  |  |  |  |  |  |  |  |  |
| Not at all | 7224 (93.8) | 6697 (92.7) | 527 (7.3) | <0.001 | 281 (53.2) | 247 (46.8) | 0.019 | 10002 (73.5) | 6380 (63.8) | 3622 (36.2) | <0.001 | 1606 (44.4) | 2015 (55.6) | <0.001 |
| Less than once a week | 56 (0.7) | 39 (69.0) | 17 (30.4) |  | 7 (43.8) | 9 (56.2) |  | 517 (3.8) | 169 (32.7) | 348 (67.3) |  | 143 (41.1) | 205 (58.9) |  |
| At least once a week | 122 (1.6) | 83 (68.0) | 39 (32.0) |  | 16 (41.0) | 23 (59.0) |  | 1072 (7.9) | 325 (30.3) | 748 (69.7) |  | 292 (39.0) | 456 (61.0) |  |
| Almost every day | 303 (3.9) | 158 (52.1) | 145 (47.9) |  | 50 (34.2) | 96 (65.8) |  | 2025 (14.9) | 462 (22.8) | 1563 (77.2) |  | 490 (31.3) | 1073 (68.7) |  |

<sup>1</sup>DHS 2017-2018<sup>3</sup>Percentage aware of cervical cancer<sup>2</sup>DHS 2018
\*Wald test of association

### Awareness of cervical cancer and cervical screening

Fig 2 shows the awareness of cervical cancer and cervical screening among women. Only 9.5% of women in Benin were aware of cervical cancer but 46.1% were aware of cervical cancer in Cameroon. Around half of the women who were aware of cervical cancer in both countries were aware of cervical screening, Benin (51.4%) and Cameroon (59.7%).

**Fig 2.**
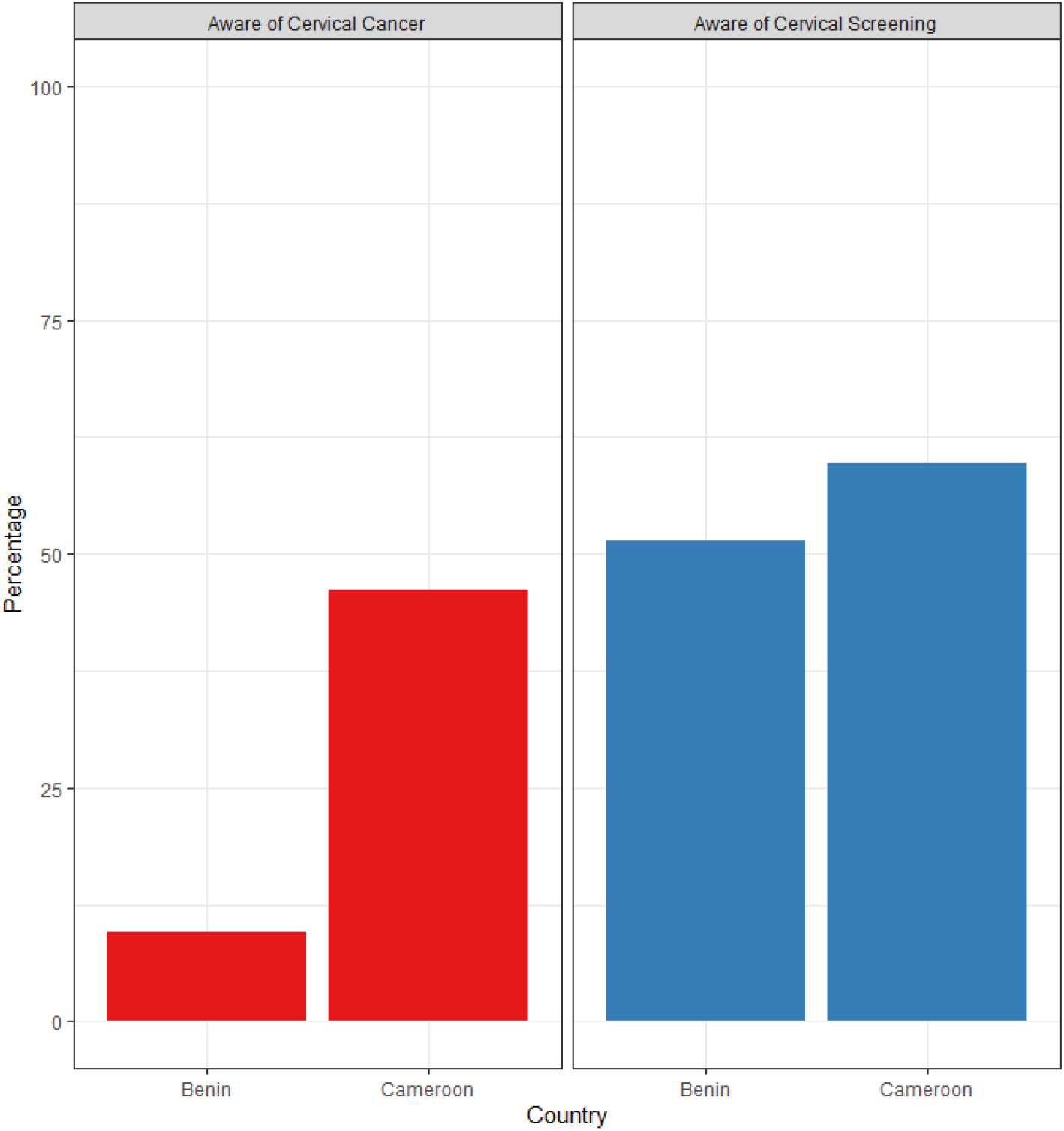
Awareness of cervical cancer and cervical screening among women of reproductive age in Benin and Cameroon.

#### Awareness of Cervical Cancer (Table 1)

In Benin awareness varied slightly by age but was very low (less than 11%), in Cameroon at least half of the women aged 25 and above were aware of cervical cancer.

Awareness was higher among women in the urban areas in Benin (14.0%) and Cameroon (57.9%). Women who had never been in a union in Benin (10.5%) and Cameroon (47.9%) were more aware of cervical cancer than women who were currently/formerly married. Women with higher education in Benin (53.0%) and Cameroon (82.2%) were more aware of cervical cancer. Similar patterns were seen in the wealth quintiles in both countries, women in the richest wealth quintile had the highest proportion of awareness in Benin (20.8%) and Cameroon (71.0%). Christian women were more aware of cervical cancer in Cameroon (54.4%) than women following other religious faiths. Professional women were more aware of cervical cancer in both countries. Women who watch television (20.2%, 63.8%) and listen to the radio (14.0%, 64.2%) at least once a week and women who use the internet almost every day (47.9%, 77.2%) were more aware of cervical cancer in Benin and Cameroon respectively (Table 2). Overall, awareness of cervical cancer in Benin was very low compared with Cameroon.

#### Awareness of cervical screening among those aware of cervical cancer (Table 1)

In both countries, awareness of cervical screening slightly varied across the different age groups. Women aged 25-34 years in Benin (58.4%) and women aged 45+ years in Cameroon (66.0%) had the highest level of awareness of cervical screening. Awareness of screening was higher in women in the urban areas of Benin (54.5%) and Cameroon (62.0%). Currently/formerly married women were more aware of screening than those who were never in a union in Benin (53.4%) and Cameroon (62.3%). Awareness of cervical screening increased with education level in both countries, women with higher education had greater awareness of cervical screening in Benin (71.6%) and Cameroon (74.4%). Awareness of cervical screening had no clear pattern of association with the wealth quintiles, but in both countries, women in the richest wealth quintile had the highest proportion of awareness in Benin (56.6%) and Cameroon (65.8%). Christian women in Benin (52.7%) were more aware of cervical screening but in Cameroon, women with other religions (68.9%) were more aware of screening. Professional women in both countries were more aware of cervical screening in Benin (63.5%) and Cameroon (65.9%).

Women in both countries who read newspapers/magazines, watched television, and listened to the radio at least once a week were more aware of cervical screening. The highest awareness was seen in women who read newspapers/magazines in Benin (65.7%) and Cameroon (67.7%). Women who used the internet almost every day in the last month were more aware of cervical screening in Benin (65.8%) and Cameroon (68.7%).

### Logistic regression of factors associated with awareness of cervical cancer

Univariate logistic regressions were conducted to provide a direct comparison with the multivariate logistic regressions (Table 3). In line with the results of the chi-square analysis (Tables 1 and 2), all the demographic characteristics and media use in both countries were significantly associated with awareness of cervical cancer except marital status. The results of the univariate analysis are shown in S1 Table. The multivariate logistic regression included the following variables in the adjusted model: age, residential status, marital status, education, wealth quintiles, religion, occupation, and frequencies of reading newspapers or magazines, watching television, listening to the radio, and using the internet in the last month.

**Table 3:**
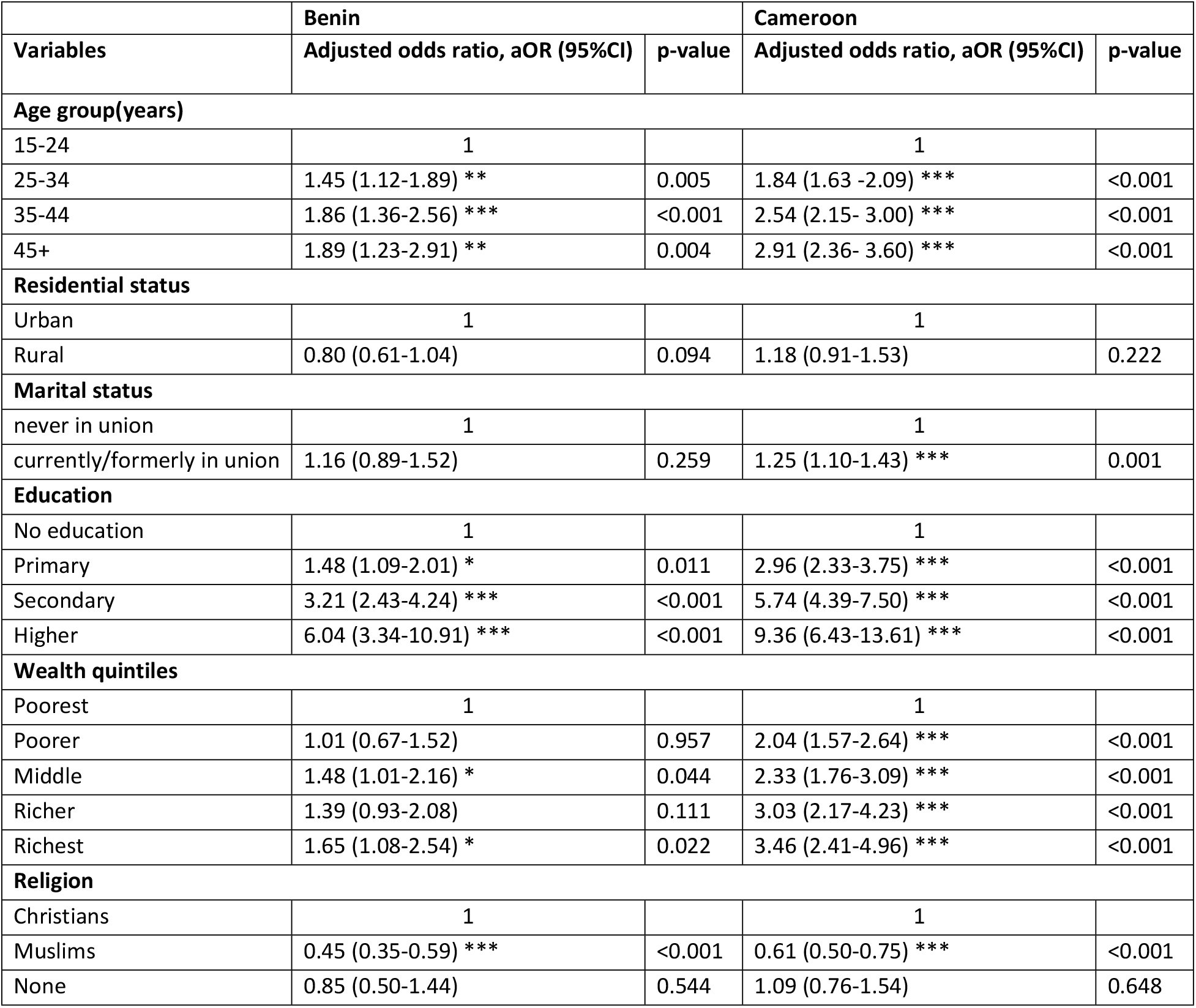
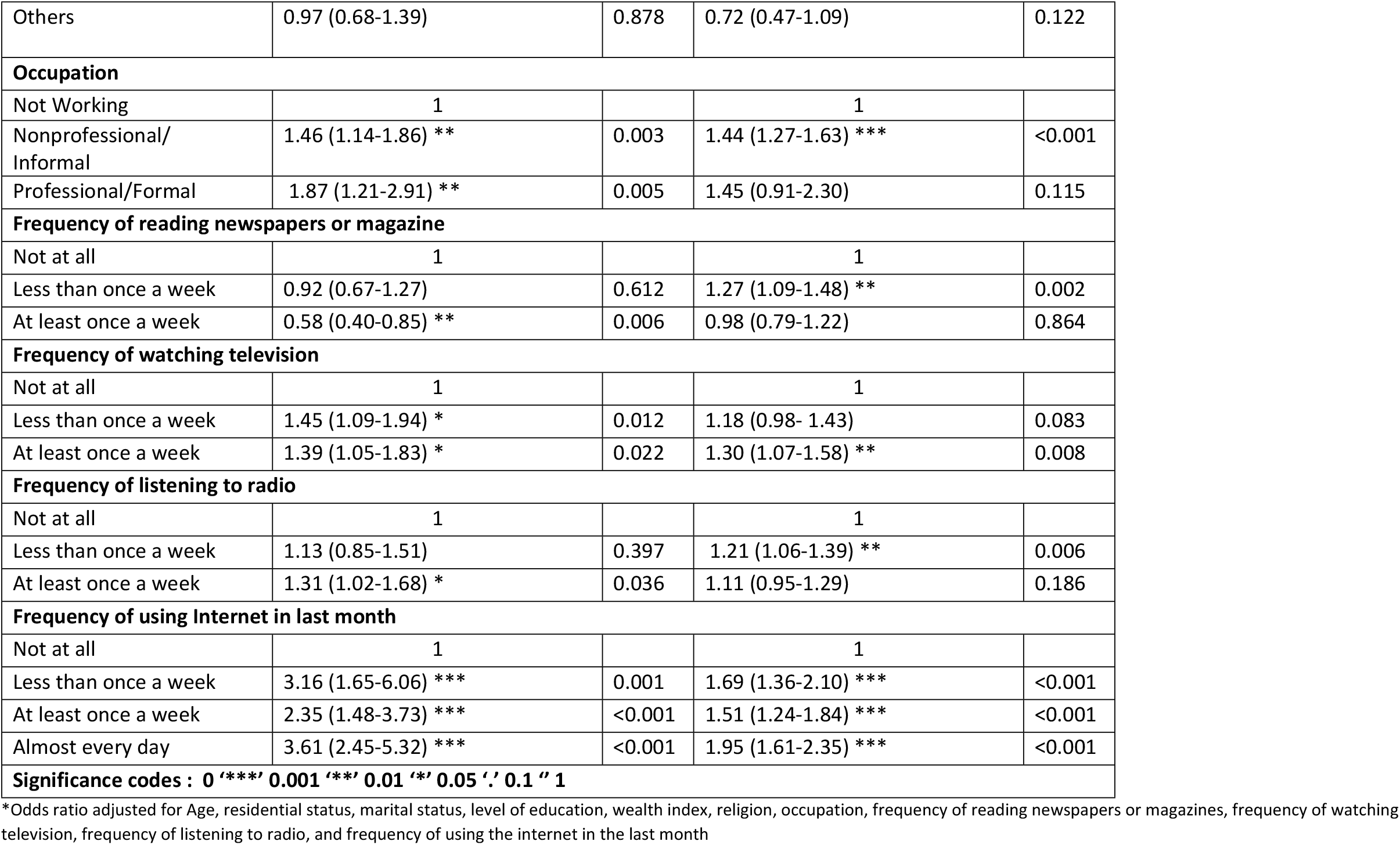
Demographic characteristics and media use associated with awareness of cervical cancer among women 15-49 years in Benin and Cameroon.

In Benin, the odds of awareness of cervical cancer significantly increased from women aged 25-34 years (adjusted odds ratio, aOR 1.45, 95% CI 1.12-1.89, p= 0.005) to women aged 45+ years (aOR 1.89, 95% CI 1.23-2.91, p= 0.004) compared with women aged 15-24 (Table 3). The awareness of cervical cancer increased as the education levels increased in Benin, the odds were approximately 6 times higher (95% CI 3.34-10.91, p= <0.001) in women with higher education than in women without education in the adjusted model. The middle and richest wealth quintiles in the adjusted model had significantly increased odds of awareness of cervical cancer (Table 3). Muslims in the adjusted model in Benin had significantly reduced odds of awareness (aOR 0.45, 95% CI 0.35-0.59, p= <0.001) than Christians. Being a professional woman in Benin increased the odds of awareness of cervical cancer (aOR 1.87, 95% CI 1.21-2.91, p=0.005). There was a significantly reduced odds ratio among women who read newspapers or magazines at least once a week (aOR 0.58, 95% CI 0.40-0.85, p= 0.006). Benin women who watched television less than once a week had the highest odds (aOR 1.45, 95% CI 1.09-1.94, p= 0.022) while women who listened to the radio at least once a week had the highest odds (aOR 1.31, 95% CI 1.02-1.68, p= 0.036) than those who do not do that at all. There was a reduction in the odds among women who use the internet at least once a week, but the odds were significantly increased in women who used the internet almost every day, (aOR 3.61, 95% CI 2.45-5.32, p= <0.001) (Table 3).

In Cameroon, the odds of awareness of cervical cancer increased as age increased in the adjusted models (Table 3). The odds were highest (aOR 2.91, 95% CI 2.36-3.60, p= <0.001) among women aged 45+ years compared with women aged 15-24 years. Women who were currently /formerly married had a higher odd (aOR 1.25, 95% CI 1.10-1.43, p= 0.001) than the unmarried in the adjusted model. Awareness of cervical cancer was approximately nine times higher (aOR 95% CI 6.43-13.61, p= <0.001) among women with higher education than women with no education. Similarly, awareness significantly increased as the wealth index increased in the adjusted model, approximately four times higher (aOR 95% CI 2.41-4.96, p= <0.001) in the richest wealth quintile compared to women in the poorest wealth quintile. Muslims in the adjusted model had reduced odds of cervical cancer awareness (aOR 0.61, 95% CI 0.50-0.75, p= <0.001) than Christians. The non-professional women had significantly increased odds in awareness (aOR 1.44 95% CI 1.27-1.63, p= <0.001) compared with the not working women. There were significantly increased odds of cervical cancer awareness in Cameroon among women who read newspapers/magazines (aOR 1.27, 95% CI 1.09-1.48, p= 0.002) and listened to the radio (aOR 1.21, 95% CI 1.06-1.39, p= 0.006) less than once a week than those who did not. Also, among those that watch television at least once a week the odds increased (aOR 1.30, 95% CI 1.07-1.58, p= 0.008). The odds were highest in those that used the internet almost every day (aOR 1.95, 95% CI 1.61-2.35, p= <0.001).

In summary, older age, higher education levels, and frequent use of the internet in the last month were the factors with a clear significant association with increased cervical cancer awareness in the two countries (Table 3). Additionally in Cameroon, the wealth index had a clear significant association with awareness of cervical cancer.

### Logistic regression analysis of factors associated with awareness of cervical cancer screening

In the chi-square analysis (Tables 1 and 2); age, education, wealth index, occupation, and media usage (television, radio, internet) were associated with awareness of cervical cancer screening in Benin whereas all the demographic characteristics and media use were associated with awareness of cervical cancer screening in Cameroon. The results of the univariate analysis are in the S2 Table.

In Benin, the odds of cervical screening awareness were significantly higher (aOR 1.98, 95% CI 1.20-3.27, p= 0.007) among women aged 25-34 years than aged 15-24 years in the adjusted model (Table 4). Women with higher education had four-fold increased odds of cervical screening awareness (aOR 4.42,95% CI 1.85-10.55, p= 0.001) compared with women with no education in the adjusted model. Women who listened to the radio at least once a week in Benin had significantly increased odds of cervical screening awareness (aOR 1.73, 95% CI 1.04-2.88, p= 0.034) in the adjusted model.

**Table 4.** Demographic characteristics and media use associated with awareness of cervical cancer screening among women 15-49 years in Benin and Cameroon.

|  | **Benin** |  | **Cameroon** |  |
| --- | --- | --- | --- | --- |
| **Variables** | **Adjusted odds ratio (95%CI)** | **p-value** | **Adjusted odds ratio (95%CI)** | **p-value** |
| **Age group(years)** |  |  |  |  |
| 15-24 | 1 |  | 1 |  |
| 25-34 | 1.98 (1.20-3.27) ** | 0.007 | 1.31 (1.11-1.54) ** | 0.002 |
| 35-44 | 1.72 (0.93-3.18) | 0.084 | 1.66 (1.38-2.00) *** | <0.001 |
| 45+ | 1.81 (0.93-3.54) | 0.08 | 1.75 (1.33-2.29) *** | <0.001 |
| **Residential status** |  |  |  |  |
| Urban | 1 |  | 1 |  |
| Rural | 0.92 (0.56-1.51) | 0.754 | 0.96 (0.77-1.19) | 0.686 |
| **Marital status** |  |  |  |  |
| never in union | 1 |  | 1 |  |
| currently/formerly in union | 1.66 (0.98-2.80) | 0.059 | 1.29 (1.09-1.53) ** | 0.004 |
| **Education** |  |  |  |  |
| no education | 1 |  | 1 |  |
| primary | 1.40 (0.78-2.54) | 0.26 | 1.41 (0.96-2.08) | 0.082 |
| secondary | 2.22 (1.22-4.06) ** | 0.01 | 1.53 (1.02-2.30) * | 0.041 |
| higher | 4.42 (1.85-10.55) *** | 0.001 | 2.69 (1.59-4.55) *** | <0.001 |
| **Wealth quintiles** |  |  |  |  |
| Poorest | 1 |  | 1 |  |
| Poorer | 1.43 (0.62-3.29) | 0.396 | 0.84 (0.52-1.35) | 0.461 |
| Middle | 0.61 (0.28-1.33) | 0.215 | 0.78 (0.49-1.26) | 0.314 |
| Richer | 0.96 (0.47-1.99) | 0.92 | 0.89 (0.53-1.48) | 0.642 |
| Richest | 0.84 (0.38-1.86) | 0.671 | 0.96 (0.57-1.62) | 0.873 |
| **Religion** |  |  |  |  |
| Christians | 1 |  | 1 |  |
| Muslims | 0.98 (0.61-1.58) | 0.93 | 0.95 (0.75-1.21) | 0.693 |
| None | 1.08 (0.40-2.91) | 0.886 | 0.73 (0.45-1.18) | 0.198 |
| Others | 0.97 (0.48-1.97) | 0.941 | 1.43 (0.80-2.57) | 0.23 |
| **Occupation** |  |  |  |  |
| Not Working | 1 |  | 1 |  |
| Nonprofessional/Informal | 0.77 (0.48-1.24) | 0.287 | 1.02 (0.88-1.19) | 0.752 |
| Professional/Formal | 0.72 (0.37-1.39) | 0.321 | 0.87 (0.60-1.24) | 0.433 |
| **Frequency of reading newspapers or magazine** |  |  |  |  |
| Not at all | 1 |  | 1 |  |
| Less than once a week | 0.68 (0.41-1.13) | 0.132 | 0.98 (0.84-1.13) | 0.75 |
| At least once a week | 1.05 (0.51-2.13) | 0.901 | 1.06 (0.83-1.36) | 0.629 |
| **Frequency of watching television** |  |  |  |  |
| Not at all | 1 |  | 1 |  |
| Less than once a week | 0.78 (0.45-1.34) | 0.362 | 0.83 (0.66-1.05) | 0.118 |
| At least once a week | 0.96 (0.53-1.72) | 0.884 | 1.04 (0.84-1.27) | 0.741 |
| **Frequency of listening to radio** |  |  |  |  |
| Not at all | 1 |  | 1 |  |
| Less than once a week | 1.15 (0.64-2.05) | 0.637 | 0.98 (0.84-1.15) | 0.806 |
| At least once a week | 1.73 (1.04-2.88) * | 0.034 | 1.01 (0.84-1.23) | 0.88 |
| **Frequency of using Internet in last month** |  |  |  |  |
| Not at all | 1 |  | 1 |  |
| Less than once a week | 1.27 (0.43-3.77) | 0.669 | 1.13 (0.86-1.48) | 0.381 |
| At least once a week | 1.32 (0.62-2.78) | 0.471 | 1.08 (0.88-1.32) | 0.448 |
| Almost every day | 1.39 (0.76-2.54) | 0.287 | 1.35 (1.10-1.67) ** | 0.005 |
| **Significance codes : 0 '\*\*\*' 0.001 '\*\*' 0.01 '\*' 0.05 '.' 0.1 ' ' 1** |  |  |  |  |

\*Odds ratio adjusted for Age, residential status, marital status, level of education, wealth index, religion, occupation, frequency of reading newspapers or magazines, frequency of watching television, frequency of listening to radio, and frequency of using the internet in the last month

In Cameroon, the odds of awareness of cervical screening significantly increased as age increased (Table 4). Women aged 45+ years had an increased odds of awareness compared with women aged 15-24 years (aOR 1.75, 95% CI 1.33-2.29, p= <0.001). Women who were currently/formerly married had increased odds of cervical screening awareness (aOR 1.29, 95% CI 1.09-2.53, p= 0.004) than women who had never been in a union in the adjusted model. The odds of cervical screening awareness significantly increased in women with secondary education (aOR 1.53, 95% CI 1.02-2.30, p= 0.041) to approximately three times higher (aOR 2.69, 95% CI 1.59-4.55, p= <0.001) in women with higher education in the adjusted model. Using the internet almost every day in the last month significantly increased the odds of cervical screening awareness (aOR 1.35, 95% CI 1.10-1.67, p= 0.005) in Cameroon.

In summary, older age, a higher level of educational achievement, and frequency of use of radio and the internet were significantly associated with the awareness of cervical screening.

## Discussion

This study assessed the relationship between sociodemographic factors and awareness of cervical cancer and cervical screening in Benin and Cameroon, using the nationally representative DHS sample. Awareness of cervical cancer was significantly associated with older age, higher educational achievement, and frequent use of the internet in the last month in both countries, and with greater wealth in Cameroon. Awareness of cervical cancer screening was highest among women aged 25-34 years in Benin, women aged 45+ in Cameroon, and women with higher education in both countries. Age, education, and frequency of use of radio and the internet were also significantly associated with the awareness of cervical screening.

Awareness of cervical cancer varies across developing countries. In our study, awareness of cervical cancer was as low as 9.5% in Benin and 46.1% in Cameroon. In the Tanzania HIV and Malaria Indicators Survey, only 30.9% of women aged 15-49 years were unaware of cervical cancer, implying that more than 60% of the women were aware of cervical cancer in Tanzania (25). Predictors of cervical cancer awareness in that study included having more than secondary education, being affluent, having 1-4 children, and being aged 30-44 years (25). The risk of cervical cancer is increased in women with HIV; hence it is the most frequently detected cancer among women with HIV (26). Tanzania is one of the sub-Saharan African countries with higher prevalence of HIV compared to the prevalence of HIV worldwide (27). Therefore, women who participated in the Tanzania survey (25), could be more aware of cervical cancer because of the increased risk of cervical cancer associated with HIV. In a DHS analysis on the prevalence of awareness of cervical cancer in Zimbabwe (DHS 2015) and Benin (DHS 2017-18), awareness of cervical cancer in Zimbabwe was as high as 79.2%, much higher than our study and awareness in Benin (10.2%) was similar to our study (28). It appears the whole sample of women interviewed during the survey were used but, in our study, we analysed a sub-sample of women who were asked questions about cervical cancer. This is the reason a direct comparison cannot be made between our study and theirs. A population-based cross-sectional study of 1,090 women in Bangladesh, revealed an awareness of cervical cancer of 45.2% among women 15 to 75 years (29). This result is close to the prevalence in Cameroon in our study; although, the upper age limit in our study was only 49 years. Marital status, literacy, residential status, and socio-economic status were found to be associated with the level of knowledge of the women in the Bangladesh study (29). A nationally representative household survey that considered awareness of specific cervical screening tests such as smear tests among women also found low levels of awareness in Nepal (13%) (30), and Indonesia (20%)(31). In our study, only women who were aware of cervical cancer were asked about awareness of cervical screening. Hence the reason why around half of the women in Benin (51.4%) and Cameroon (59.7%) were aware of cervical screening. Our study did not consider if the women were aware of the different methods of cervical screening. Cervical cancer screening programmes in most African countries are less than the WHO target of 70% coverage, only 20% of the WHO countries in the African region have a national cervical cancer screening programme which is mainly opportunistic (32). As of 2021, Benin and Cameroon had no national HPV vaccination programme and no screening programme for cervical cancer (7, 8). The possibility that women will be aware of cervical cancer and screening is likely to be lower in countries without a national screening programme. There is a need to increase the level of awareness of cervical cancer and screening among women in Benin and Cameroon through stakeholder involvement and the establishment of an implementable national cervical screening programme.

The significant determinants of awareness of cervical cancer from our study included age, education, and frequency of using the internet in the last month in both countries and the wealth quintiles in Cameroon. Studies show that as a woman’s age increases she will possibly be more aware of the health issues that are associated with her reproductive health (28). Alongside advanced maternal age, studies show that having a formal education, using the internet, and having a professional/technical/managerial occupation significantly increased the odds of awareness of cervical cancer after adjusting for other confounders (28). Like our study, women aged over 45 years had the highest odds of cervical cancer awareness (28). Our study differs from Barrow et al (28) in the forms of media used by women to get their information on health issues. In our study, women using the internet in the last month in both countries had better awareness of cervical cancer. Some previous studies did not specify the media use type, although some identified television as the best means of cervical cancer information delivery (33, 34). Online social media platforms have been identified as a means for public health promotion but there is limited research on the effectiveness of social media in improving awareness of health topics (33). Inadequate information about cervical cancer could result in poor knowledge about the disease (34). It appears that studies agree that women get their health information from various forms of media, therefore, media use could improve awareness of cervical cancer.

Awareness of cervical cancer differed by wealth quintiles in Cameroon. In Ethiopia (35) women with a higher average monthly income had better awareness of cervical cancer while in Kenya (36) regional disparities in cancer awareness and screening utilization services are higher among the vulnerable regions with higher rural-urban wealth inequalities. Therefore, in most low-resource settings, age, education level, residential status, and socioeconomic status predict awareness of either cervical cancer or screening (25, 28, 29, 31). There is a need to increase awareness among women of low socio-economic status to reduce the health inequalities that exist in their access to cervical cancer screening.

In Benin, an integrated strategic plan was developed to combat non-communicable diseases including cervical cancer (37). A similar plan was developed in Cameroon in 2020 for cancer control (16). As part of its primary prevention strategies, Cameroon employed awareness creation on special days and events, and in 2015 it completed a pilot phase to introduce the HPV vaccine for girls aged 9-13 years but the vaccine has not been added to the Expanded Program on Immunisation (EPI) since the pilot study (16). The cervical screening programmes available in Cameroon are those organised by some public hospitals, faith-based organisations hospitals, National Committee for the Fight against Cancer (NACFAC), and Civil Society Organisations (CSOs) (16). In both Benin and Cameroon, fewer than 1 in 10 women have had a cervical screening in the last 5 years (7, 8). Awareness creation through active stakeholder participation is needed in both countries. Developing communication and behaviour change materials during the opportunistic programme may help increase awareness in women.

### Study Limitations

Our study used a large nationally representative sample, it is a cross-sectional study and causality cannot be established but the results are generalisable to women of reproductive age groups in the countries. We had results for only a small sub-sample, so findings may be subject to sampling bias. Data were collected through interviewer-assisted self-reporting and thus subject to recall bias. We could not adjust for unmeasured confounders; therefore, we cannot rule out the role of residual confounding.

## Conclusions

Data from this study will contribute to the existing literature on awareness of cervical cancer and screening, educate cervical cancer stakeholders, and inform future intervention research to help increase awareness of cervical cancer. Awareness of cervical cancer and screening in Benin and Cameroon is very low. There is a need to strengthen awareness programmes through stakeholder engagement and public enlightenment. Media use, such as radio listening and internet use, may have a potential role in improving public enlightenment. In both countries, stakeholders need to implement the strategic plan for cervical cancer control and intensify awareness creation. Targeting younger women with age-appropriate cervical cancer information will help tackle the low level of awareness among young women. The inequality in awareness of cervical cancer and screening that exist between the poor and the rich in Cameroon needs to be tackled by investing in the education of the poor and ending residential segregation of access to health care. This will possibly allow free access to the required information on cervical cancer and screening.

## Data Availability

All the data are made available on authorisation by the DHS repository (http://dhsprogram.com/).

## Supporting Information

**S1 Table**. Univariate logistic regression of the demographic characteristics and media use associated with awareness of cervical cancer among women 15-49 years in Benin and Cameroon

**S2 Table**. Univariate logistic regression of the demographic characteristics and media use associated with awareness of cervical cancer screening among women 15-49 years in Benin and Cameroon

## Acknowledgments

We appreciate the women who participated in the survey and the DHS program team.

